# Work-related and Personal Factors Associated with Mental Well-being during COVID-19 Response: A Survey of Health Care and Other Workers

**DOI:** 10.1101/2020.06.09.20126722

**Authors:** Bradley A. Evanoff, Jaime R. Strickland, Ann Marie Dale, Lisa Hayibor, Emily Page, Jennifer G. Duncan, Thomas Kannampallil, Diana L. Gray

**Author notes:** Corresponding Author: Bradley A. Evanoff, M.D., Healthy Work Center, Washington University School of Medicine in St. Louis, 4523 Clayton Avenue, Campus Box 8005, St. Louis, MO 63110.

## Abstract

**Objective:** Measure the prevalence of stress, anxiety, depression, work-exhaustion, burnout, and decreased well-being among faculty and staff at a university and academic medical center during the SARS-CoV-2 pandemic, and describe work-related and personal factors associated with mental health and well-being.

**Design:** Observational cohort study conducted between April 17 and May 1, 2020 using a web-based questionnaire.

**Setting:** Medical and main campuses of a university.

**Participants:** All faculty, staff, and post-doctoral fellows.

**Exposures:** Work factors including supervisor support and exposure to high-risk clinical settings; personal factors including demographics and family/home stressors.

**Main Outcomes and Measures:** Stress, anxiety, depression, work exhaustion, burnout, and decreased well-being.

**Results:** There were 5550 respondents (overall response rate of 34.3%). 38% of faculty and 14% of staff (n=915) were providing clinical care, while 57% of faculty and 77% of staff were working from home. The prevalence of anxiety, depression, and work exhaustion were somewhat higher among clinicians than non-clinicians. Among all workers, anxiety, depression, and high work exhaustion were independently associated with community or clinical exposure to COVID-19 [Prevalence Ratios and 95% confidence intervals 1.37(1.09- 1.73), 1.28(1.03 - 1.59), and 1.24(1.13 - 1.36) respectively]. Poor family supportive behaviors by supervisors were also associated with these outcomes [1.40 (1.21 - 1.62), 1.69 (1.48 - 1.92), 1.54 (1.44 - 1.64)]. Age below 40 and a greater number of family/home stressors were also associated with poorer outcomes. Among the subset of clinicians, caring for patients with COVID-19 and work in high-risk clinical settings were additional risk factors.

**Conclusions and Implications:** Our findings suggest that the pandemic has had negative effects on mental health and well-being among both clinical and non-clinical employees. Prevention of exposure to COVID-19 and increased supervisor support are modifiable risk factors that may protect mental health and well-being.

## INTRODUCTION

The SARS-CoV-2 pandemic has created unprecedented disruption in social interactions and working conditions. Recent studies have described the effects of the pandemic on the mental health and well-being of frontline healthcare workers (HCW),^1,2^ and potential interventions to protect them.^3-5^ Although concern over health and well-being has primarily focused on frontline HCW, the pandemic has also affected working conditions in most other industries. Social and employment changes have led to concern of an impending “second pandemic” of short and long term mental health issues, ^6^ and predictions of a preventable surge of avoidable deaths from alcohol, drug use, and suicide.^7^ Few data describe the effects of the pandemic on mental health and well-being of workers outside of healthcare. Such evidence is important for developing appropriate responses to the pandemic in order to preserve health and plan for economic and social recovery.

We describe results from the EMPOWER study (**Emp**l**o**yee **W**ell-Being during **E**pidemic **R**esponse), which measured mental health and well-being among a large and diverse academic workforce, including those with and without clinical exposure to COVID-19 patients. The goals of the study were to measure the prevalence of, stress, anxiety, depression, work exhaustion, burnout, and decreased mental well-being among faculty and staff at a university and its academic medical center during the SARS-CoV-2 pandemic; to compare mental health and well-being between clinical workers who were or were not caring for COVID-19 patients; and to identify other modifiable workplace and personal risk factors associated with mental health and well-being.

## METHODS

### Study Design and Participants

We conducted a web-based survey of all benefits-eligible university employees (faculty, staff, and post- doctoral scholars) at Washington University in St. Louis, a private university with a large academic medical center where attending physicians and clinical staff are university employees. A separate survey was sent to physician trainees (residents and clinical fellows) and is not included in this report. An email invitation to participate was sent to all benefits-eligible employees on April 17, 2020, with a clickable link to a voluntary, anonymous online survey. A single reminder email was sent ten days later. The survey period spanned 4 -5 weeks after the university enacted work at home plans. The study was approved by the institutional review board of Washington University in St. Louis.

### Survey Instrument

The survey was designed to take less than 10 minutes to complete (Supplementary Materials). Demographic questions included age, race, household income, children, dependents, and other adults living at home, and work status of partner. Questions about work included current work status (onsite work involving clinical care, onsite work not involving clinical care, working from home, or not working). Those doing onsite work in clinical care were asked about clinical setting, and if they had cared for patients with COVID-19. All participants were asked if they or a member of their household had received a medical diagnosis or a positive test for COVID-19 or if they had been exposed to someone with COVID-19.

The questionnaire also included three questions from the Family Supportive Supervisor Behavior Short-Form (FSSB-SF),^8^ which measures supervisor behaviors supportive of family roles (“Your supervisor makes you feel comfortable talking to him/her about your conflicts between work and non-work; Your supervisor demonstrates effective behaviors in how to juggle work and non-work issues; Your supervisor works effectively with employees to creatively solve conflicts between work and non-work.”) We used the mean value of these three responses as the supervisor support variable. We also asked about 8 potential family/home stressors related to the pandemic (childcare, home schooling, caring for elderly relatives, having access to food and other essential ies, being infected, friends and family being infected, keeping your job, and personal finances). These questions were asked in the format “Currently how stressed are you about…?” in a 5-point scale from “not at all” to “extremely” stressed. The number of stressors reported by each individual as “somewhat” to “extremely” were totaled to create a composite stress score (range 0-8).

### Outcome Measures

Study outcomes included stress, anxiety, and depression as measured by the DASS-21,^9^ burnout and work exhaustion as measured by the Professional Fulfillment Index (PFI),^10^ and changes in well-being.^11^ The DASS-21 is a validated instrument with scales that correlate well with other measures of depression, anxiety, and stress. Due to the PFI questionnaire structure, burnout was only assessed among HCW. Self-reported changes in well-being comparing current to pre-pandemic status were assessed in five domains (overall, financial, physical, mental, and social) by the question “To what extent have COVID-19-related work/life changes impacted your well-being” using a four-point scale from “much worse” to “much better/somewhat better. “

### Statistical Analyses

We contrasted the proportions or means of outcomes between faculty and staff and those in different clinical settings, and conducted univariable and multivariable Poisson regression with robust sandwich estimators to examine personal and work factors associated with six mental health and well-being outcomes described above: stress, anxiety, depression, burnout, work exhaustion, and changes in well-being.

In conducting these analyses, we selected a priori ten potential personal and work factors as independent variables for multivariate analysis (supervisor support, clinical work, staff [vs. faculty or post-doc], exposure to people [or patients for clinicians] with a diagnosis of COVID-19, age, sex, race, annual household income, children under 18 years living at home, and composite stressor count). Results were expressed as Prevalence Ratios (PR) with 95% confidence intervals (CI). Independent variables were dichotomized at the median scores or at relevant cut-points for ordinal variables. We categorized race and ethnicity as “under-represented groups” (those identifying as Black/African American, Native American, Hawaiian/Pacific Islander or Hispanic) and as “other.” Significance level was set at 0.05 and hypothesis tests were 2-sided. All analyses were performed with R statistical software version 4.0.0^12^ and R studio version 1.2.504.^13^

### Patient and Public Involvement

The survey was developed in collaboration with the university human resources department and employee wellness director to ensure sensitivity to current issues and to address emerging concerns about employee wellness during the pandemic response. Initial survey results have been shared with university leaders in order to highlight mental health needs of employees. Study results are driving plans to communicate broadly with faculty, staff, and trainees to highlight mental health challenges faced by our workforce and to better publicize and encourage employees to utilize available mental health resources.

## RESULTS

16,238 email invitations were sent to university faculty, staff, and post-doctoral scholars. 5706 responses were received (Figure 1); there were 5569 unique responses after the exclusion of 137 responses with duplicate self-generated identifier allowing anonymous longitudinal follow-up. 19 surveys were dropped for missing status as faculty, staff, or post-doctoral scholar, leaving 5550 respondents for analysis (870 faculty, 4470 staff, and 210 post-docs). Overall response rate was 34.3% for unique surveys. Response rates were higher for staff than for faculty (40% vs. 19.7%)

**Figure 1:**
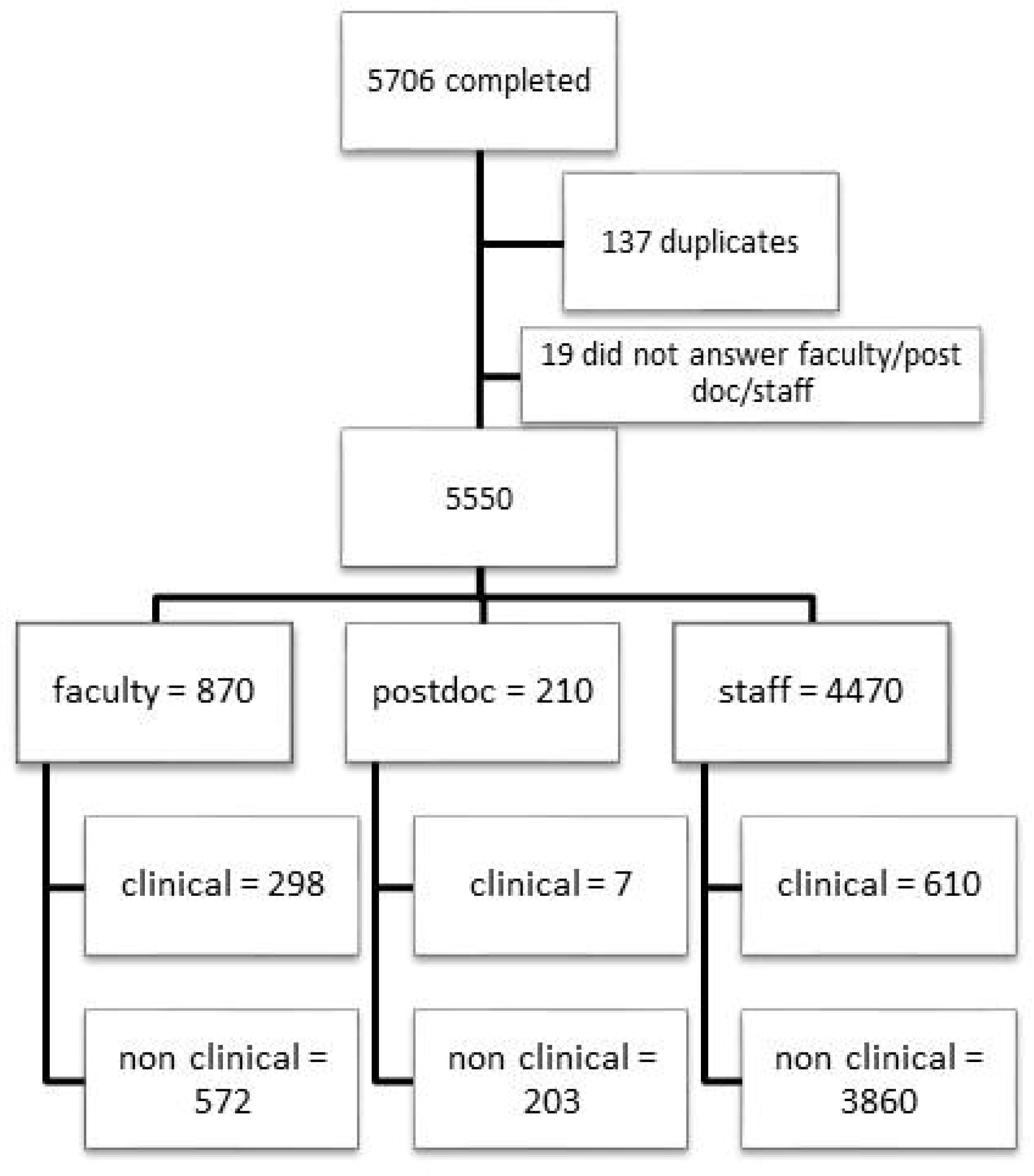
EMPOWER Study Flow Diagram

Table 1 compares demographics, work factors, and outcomes between faculty, staff, and post-docs. 34.3% of faculty and 13.6% of staff reported working onsite in clinical operations while a majority of the faculty (60.6%) and staff (76.5%) were working from home. Smaller numbers worked onsite in non-clinical roles and few were not working. A majority of faculty (50.4%) reported that their workload increased after the COVID-19 workplace changes, as compared to 40.4% of staff and 21% of post-docs. Overall, a majority of respondents reported being stressed (more than “a little bit”) about personal finances, keeping their jobs, and about themselves or friends or family being infected. Of those with children at home, a majority reported feeling stressed about home schooling; most of those providing care to elderly relatives reported stress about their care. Distributions of most perceived stressors were significantly different across the faculty, staff, and post-doctoral fellows, with post-doctoral fellows more frequently reporting stress about childcare, home schooling, and access to food and essential supplies. Faculty, staff, and post-doctoral fellows all reported high prevalence of worsened overall well-being (61.6%) related to COVID-19 work/life changes. Moderate to high levels of stress were reported by 13%, anxiety by 13%, depression by 15.9% and high work exhaustion by 43%.

**Table 1.**
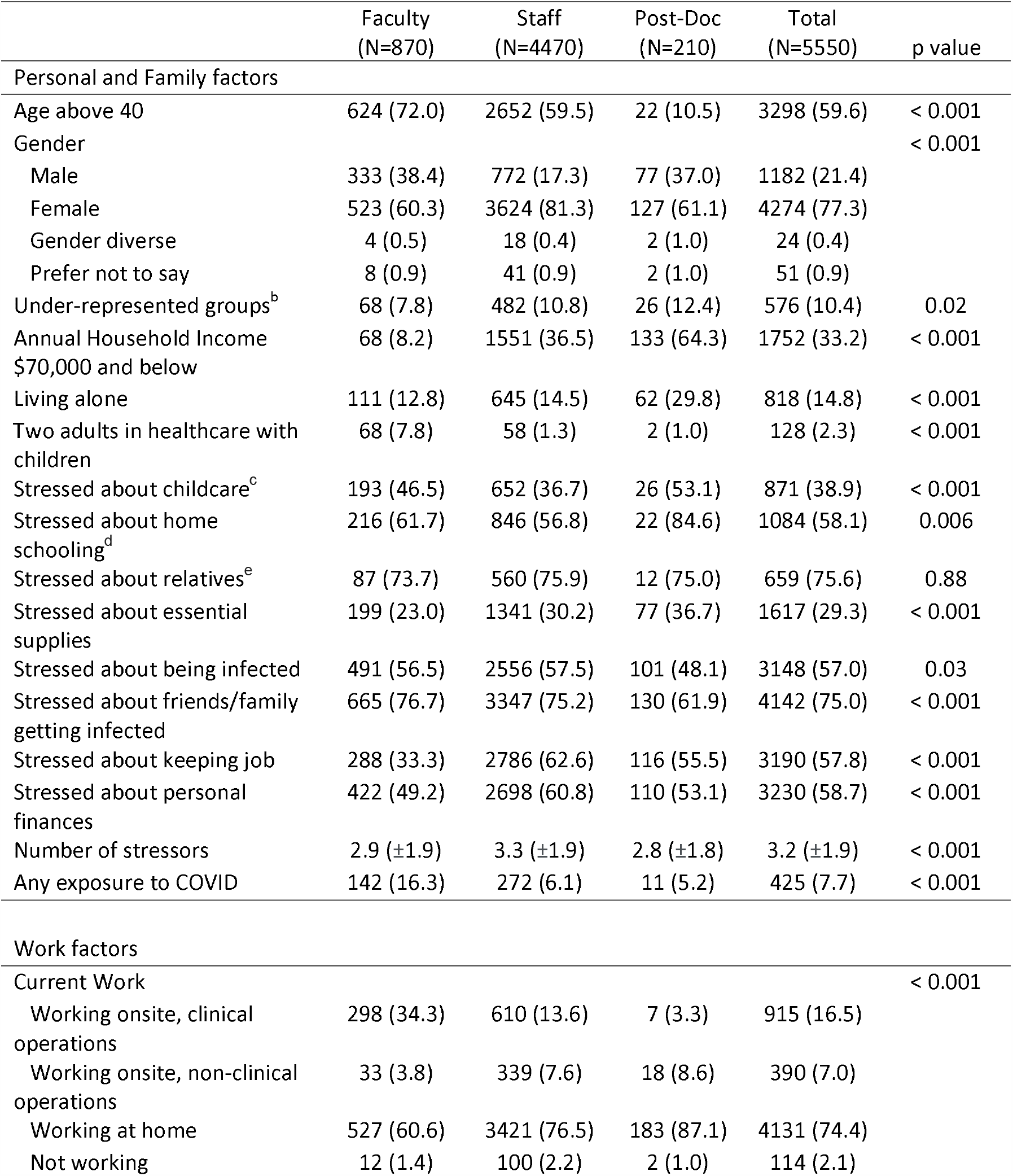
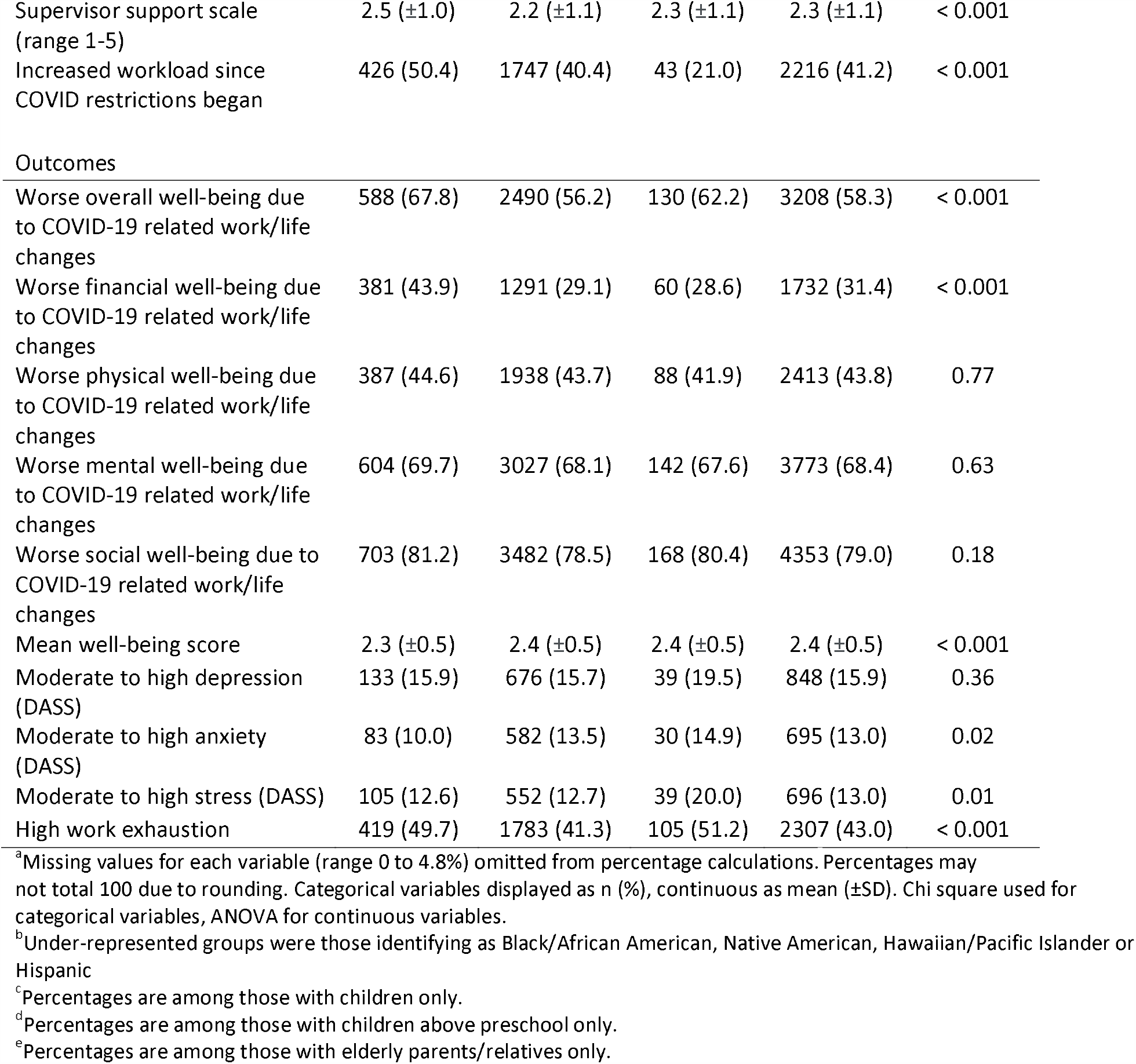
Comparison of demographics, personal factors, work factors and outcomes between faculty, staff and post-docs^a^

|  | Faculty<br>(N=870) | Staff<br>(N=4470) | Post-Doc<br>(N=210) | Total<br>(N=5550) | p value |
| --- | --- | --- | --- | --- | --- |
| Personal and Family factors |  |  |  |  |  |
| Age above 40 | 624 (72.0) | 2652 (59.5) | 22 (10.5) | 3298 (59.6) | < 0.001 |
| Gender |  |  |  |  | < 0.001 |
| Male | 333 (38.4) | 772 (17.3) | 77 (37.0) | 1182 (21.4) |  |
| Female | 523 (60.3) | 3624 (81.3) | 127 (61.1) | 4274 (77.3) |  |
| Gender diverse | 4 (0.5) | 18 (0.4) | 2 (1.0) | 24 (0.4) |  |
| Prefer not to say | 8 (0.9) | 41 (0.9) | 2 (1.0) | 51 (0.9) |  |
| Under-represented groups<sup>b</sup> | 68 (7.8) | 482 (10.8) | 26 (12.4) | 576 (10.4) | 0.02 |
| Annual Household Income<br>\$70,000 and below | 68 (8.2) | 1551 (36.5) | 133 (64.3) | 1752 (33.2) | < 0.001 |
| Living alone | 111 (12.8) | 645 (14.5) | 62 (29.8) | 818 (14.8) | < 0.001 |
| Two adults in healthcare with<br>children | 68 (7.8) | 58 (1.3) | 2 (1.0) | 128 (2.3) | < 0.001 |
| Stressed about childcare<sup>c</sup> | 193 (46.5) | 652 (36.7) | 26 (53.1) | 871 (38.9) | < 0.001 |
| Stressed about home<br>schooling<sup>d</sup> | 216 (61.7) | 846 (56.8) | 22 (84.6) | 1084 (58.1) | 0.006 |
| Stressed about relatives<sup>e</sup> | 87 (73.7) | 560 (75.9) | 12 (75.0) | 659 (75.6) | 0.88 |
| Stressed about essential<br>supplies | 199 (23.0) | 1341 (30.2) | 77 (36.7) | 1617 (29.3) | < 0.001 |
| Stressed about being infected | 491 (56.5) | 2556 (57.5) | 101 (48.1) | 3148 (57.0) | 0.03 |
| Stressed about friends/family<br>getting infected | 665 (76.7) | 3347 (75.2) | 130 (61.9) | 4142 (75.0) | < 0.001 |
| Stressed about keeping job | 288 (33.3) | 2786 (62.6) | 116 (55.5) | 3190 (57.8) | < 0.001 |
| Stressed about personal<br>finances | 422 (49.2) | 2698 (60.8) | 110 (53.1) | 3230 (58.7) | < 0.001 |
| Number of stressors | 2.9 (±1.9) | 3.3 (±1.9) | 2.8 (±1.8) | 3.2 (±1.9) | < 0.001 |
| Any exposure to COVID | 142 (16.3) | 272 (6.1) | 11 (5.2) | 425 (7.7) | < 0.001 |
| Work factors |  |  |  |  |  |
| Current Work |  |  |  |  | < 0.001 |
| Working onsite, clinical<br>operations | 298 (34.3) | 610 (13.6) | 7 (3.3) | 915 (16.5) |  |
| Working onsite, non-clinical<br>operations | 33 (3.8) | 339 (7.6) | 18 (8.6) | 390 (7.0) |  |
| Working at home | 527 (60.6) | 3421 (76.5) | 183 (87.1) | 4131 (74.4) |  |
| Not working | 12 (1.4) | 100 (2.2) | 2 (1.0) | 114 (2.1) |  |
| Supervisor support scale (range 1-5) | 2.5 ($\pm 1.0$) | 2.2 ($\pm 1.1$) | 2.3 ($\pm 1.1$) | 2.3 ($\pm 1.1$) | < 0.001 |
| Increased workload since COVID restrictions began | 426 (50.4) | 1747 (40.4) | 43 (21.0) | 2216 (41.2) | < 0.001 |
| Outcomes |  |  |  |  |  |
| Worse overall well-being due to COVID-19 related work/life changes | 588 (67.8) | 2490 (56.2) | 130 (62.2) | 3208 (58.3) | < 0.001 |
| Worse financial well-being due to COVID-19 related work/life changes | 381 (43.9) | 1291 (29.1) | 60 (28.6) | 1732 (31.4) | < 0.001 |
| Worse physical well-being due to COVID-19 related work/life changes | 387 (44.6) | 1938 (43.7) | 88 (41.9) | 2413 (43.8) | 0.77 |
| Worse mental well-being due to COVID-19 related work/life changes | 604 (69.7) | 3027 (68.1) | 142 (67.6) | 3773 (68.4) | 0.63 |
| Worse social well-being due to COVID-19 related work/life changes | 703 (81.2) | 3482 (78.5) | 168 (80.4) | 4353 (79.0) | 0.18 |
| Mean well-being score | 2.3 ($\pm 0.5$) | 2.4 ($\pm 0.5$) | 2.4 ($\pm 0.5$) | 2.4 ($\pm 0.5$) | < 0.001 |
| Moderate to high depression (DASS) | 133 (15.9) | 676 (15.7) | 39 (19.5) | 848 (15.9) | 0.36 |
| Moderate to high anxiety (DASS) | 83 (10.0) | 582 (13.5) | 30 (14.9) | 695 (13.0) | 0.02 |
| Moderate to high stress (DASS) | 105 (12.6) | 552 (12.7) | 39 (20.0) | 696 (13.0) | 0.01 |
| High work exhaustion | 419 (49.7) | 1783 (41.3) | 105 (51.2) | 2307 (43.0) | < 0.001 |

<sup>a</sup>Missing values for each variable (range 0 to 4.8%) omitted from percentage calculations. Percentages may not total 100 due to rounding. Categorical variables displayed as n (%), continuous as mean ( ± SD). Chi square used for categorical variables, ANOVA for continuous variables.
<sup>b</sup>Under-represented groups were those identifying as Black/African American, Native American, Hawaiian/Pacific Islander or Hispanic
<sup>c</sup>Percentages are among those with children only.
<sup>d</sup>Percentages are among those with children above preschool only.
<sup>e</sup>Percentages are among those with elderly parents/relatives only.

Multivariable analyses of associations between these outcomes and a common set of work and personal factors among all respondents showed three factors statistically significantly associated with a higher prevalence of all five outcomes (Table 2 – univariable analyses in supplemental materials): poor supervisor support, higher number of family/home stressors, and age below 40. Working onsite in clinical operations was associated with higher anxiety and lower mean well-being; being a staff member (rather than faculty or post-doc) was associated with better well-being and lower prevalence of stress and work exhaustion. Reported exposure to COVID-19 (diagnosis in self or family, or exposure to someone likely to have COVID-19) was associated with higher stress, anxiety, depression, and work exhaustion. Household income $70,000 and below was associated with higher prevalence of stress, anxiety, and depression. Women were more likely to report experiencing anxiety, work exhaustion, and decreased well-being. Unanticipated protective factors were also notable: having children at home was associated with lower prevalence of anxiety and depression, and underrepresented racial/ethnic groups were less likely to report stress, depression, or decreased well-being.

**Table 2.**
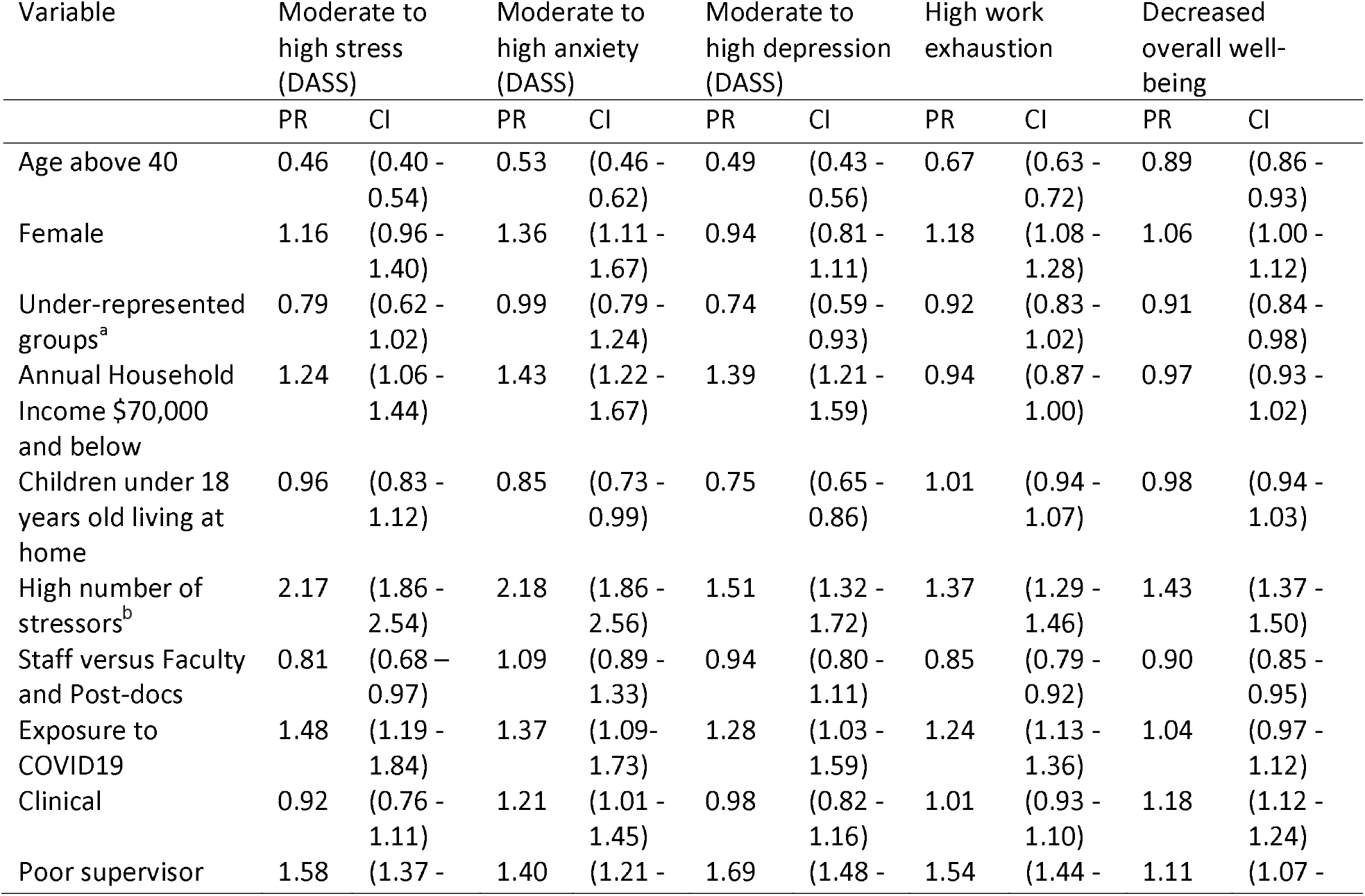
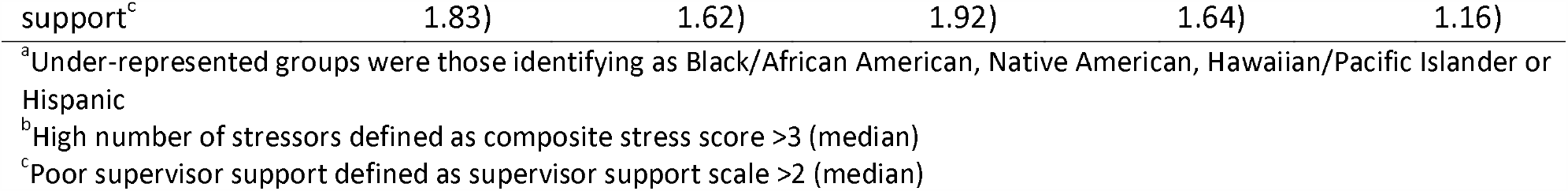
Multivariate associations between personal factors, work factors, and well-being among all participants (n = 5550, Prevalence Ratio (PR) and 95% Confidence Intervals (CI) calculated using Poisson regression models)

Comparison of outcomes between faculty and staff working in clinical settings is shown in Table 3 (univariable analyses in supplemental materials). Those working in high risk settings (Intensive Care Unit, Emergency Room, or performing procedures likely to generate respiratory aerosols) were more likely to report caring for COVID-19 patients and experiencing an increased workload since COVID-19 restrictions began, had a worse mean score on changes in well-being, and were more likely to report moderate to high stress and depression, high work exhaustion, and burnout. Multivariable analysis of faculty and staff working in clinical operations showed that caring for patients who had COVID-19 was associated with higher prevalence of stress, anxiety, burnout, and work exhaustion. (Table 4) High-risk clinical work (ICU, ED, aerosol-generating procedures) showed similar, albeit weaker associations with these outcomes in multivariable analysis (data not shown). There were no statistically significant differences between clinically active staff and faculty for any outcome. Notably, low supervisor support was strongly associated with of all mental health and well-being outcomes, and a high number of family/home stressors was associated with all outcomes except depression.

**Table 3.**
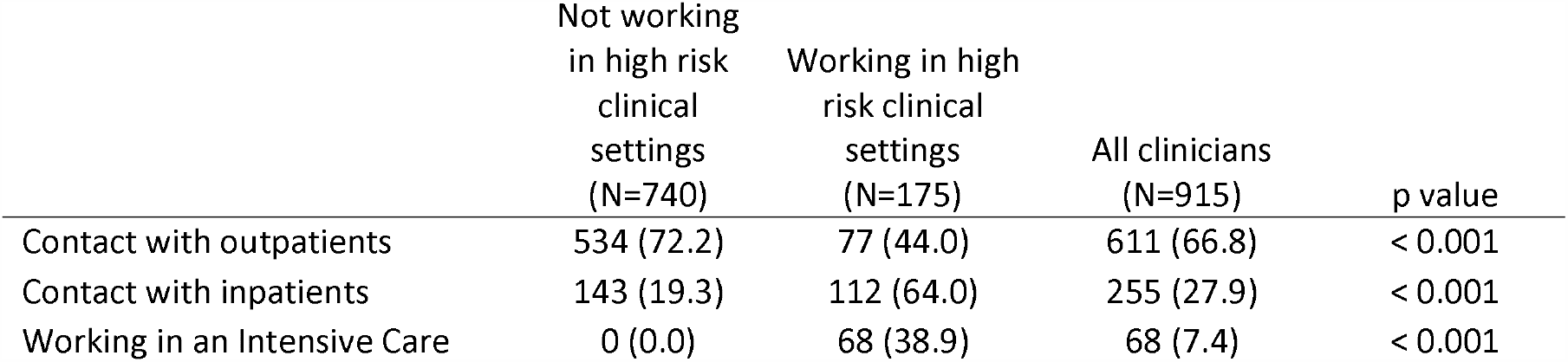
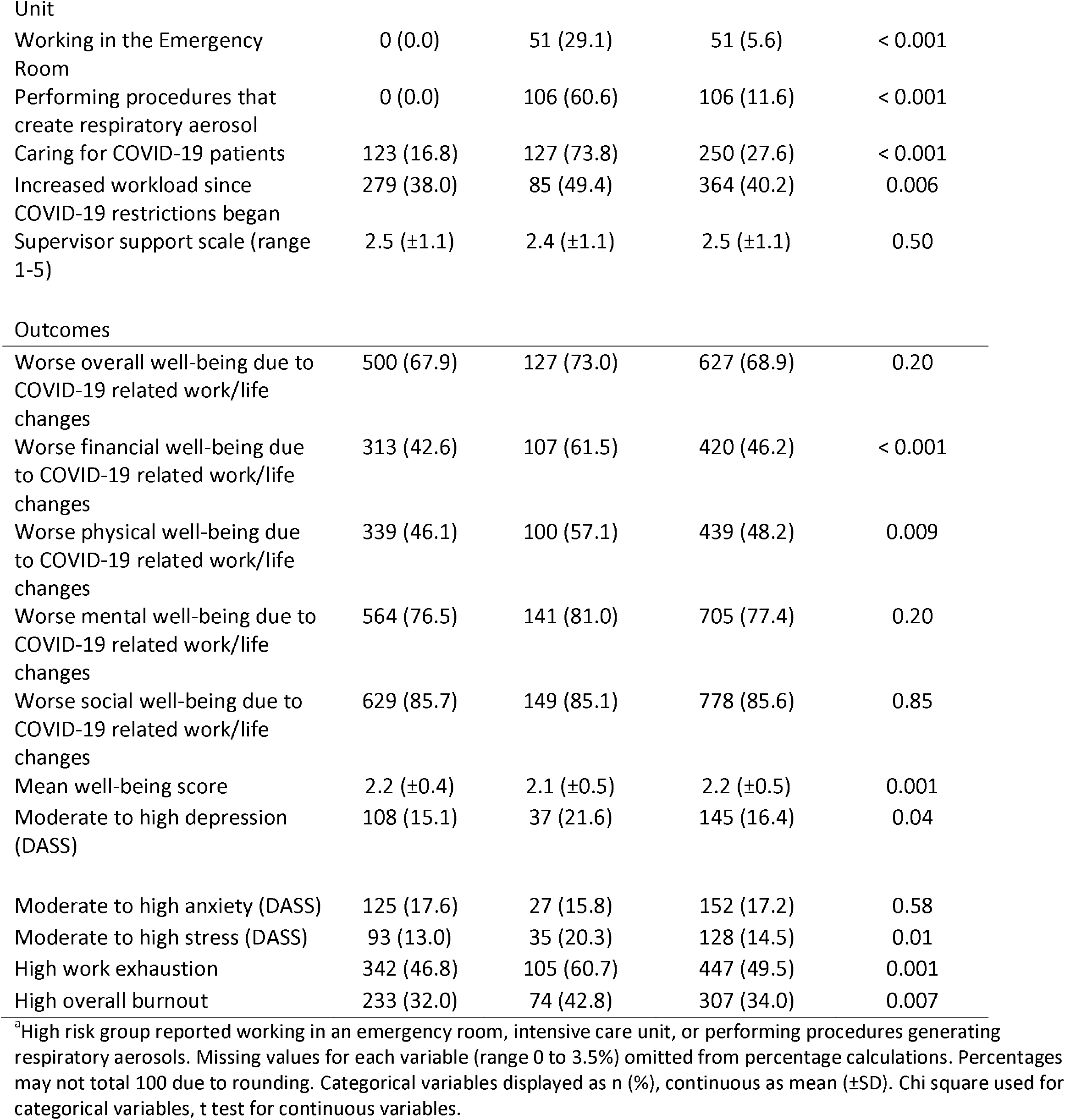
Comparison of work factors and outcomes among all clinicians and between high risk and non-high risk clinical groups^a^

**Table 4.**
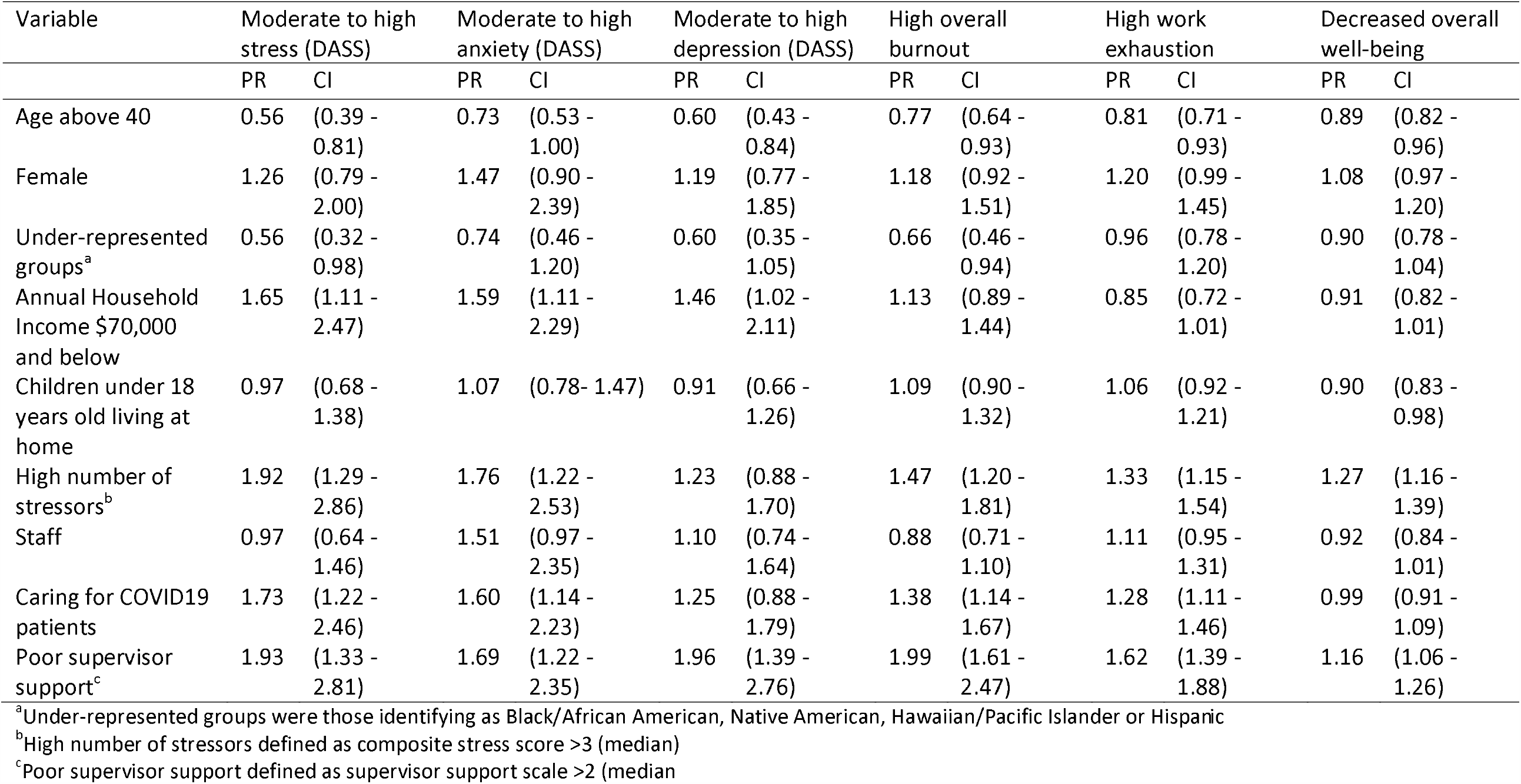
Multivariate associations between personal factors, work factors, and well-being among participants doing clinical work (n = 915, Prevalence Ratio (PR) and 95% Confidence Intervals (CI) calculated using Poisson multiple regression).

| Variable | Moderate to high stress (DASS) |  | Moderate to high anxiety (DASS) |  | Moderate to high depression (DASS) |  | High overall burnout |  | High work exhaustion |  | Decreased overall well-being |  |
| --- | --- | --- | --- | --- | --- | --- | --- | --- | --- | --- | --- | --- |
|  | PR | CI | PR | CI | PR | CI | PR | CI | PR | CI | PR | CI |
| Age above 40 | 0.56 | (0.39 - 0.81) | 0.73 | (0.53 - 1.00) | 0.60 | (0.43 - 0.84) | 0.77 | (0.64 - 0.93) | 0.81 | (0.71 - 0.93) | 0.89 | (0.82 - 0.96) |
| Female | 1.26 | (0.79 - 2.00) | 1.47 | (0.90 - 2.39) | 1.19 | (0.77 - 1.85) | 1.18 | (0.92 - 1.51) | 1.20 | (0.99 - 1.45) | 1.08 | (0.97 - 1.20) |
| Under-represented groups<sup>a</sup> | 0.56 | (0.32 - 0.98) | 0.74 | (0.46 - 1.20) | 0.60 | (0.35 - 1.05) | 0.66 | (0.46 - 0.94) | 0.96 | (0.78 - 1.20) | 0.90 | (0.78 - 1.04) |
| Annual Household Income \$70,000 and below | 1.65 | (1.11 - 2.47) | 1.59 | (1.11 - 2.29) | 1.46 | (1.02 - 2.11) | 1.13 | (0.89 - 1.44) | 0.85 | (0.72 - 1.01) | 0.91 | (0.82 - 1.01) |
| Children under 18 years old living at home | 0.97 | (0.68 - 1.38) | 1.07 | (0.78 - 1.47) | 0.91 | (0.66 - 1.26) | 1.09 | (0.90 - 1.32) | 1.06 | (0.92 - 1.21) | 0.90 | (0.83 - 0.98) |
| High number of stressors<sup>b</sup> | 1.92 | (1.29 - 2.86) | 1.76 | (1.22 - 2.53) | 1.23 | (0.88 - 1.70) | 1.47 | (1.20 - 1.81) | 1.33 | (1.15 - 1.54) | 1.27 | (1.16 - 1.39) |
| Staff | 0.97 | (0.64 - 1.46) | 1.51 | (0.97 - 2.35) | 1.10 | (0.74 - 1.64) | 0.88 | (0.71 - 1.10) | 1.11 | (0.95 - 1.31) | 0.92 | (0.84 - 1.01) |
| Caring for COVID19 patients | 1.73 | (1.22 - 2.46) | 1.60 | (1.14 - 2.23) | 1.25 | (0.88 - 1.79) | 1.38 | (1.14 - 1.67) | 1.28 | (1.11 - 1.46) | 0.99 | (0.91 - 1.09) |
| Poor supervisor support<sup>c</sup> | 1.93 | (1.33 - 2.81) | 1.69 | (1.22 - 2.35) | 1.96 | (1.39 - 2.76) | 1.99 | (1.61 - 2.47) | 1.62 | (1.39 - 1.88) | 1.16 | (1.06 - 1.26) |

<sup>a</sup>Under-represented groups were those identifying as Black/African American, Native American, Hawaiian/Pacific Islander or Hispanic
<sup>b</sup>High number of stressors defined as composite stress score >3 (median)
<sup>c</sup>Poor supervisor support defined as supervisor support scale >2 (median)

## DISCUSSION

The EMPOWER study found high prevalence of stress, anxiety, depression, work exhaustion, burnout, and worsened well-being among clinical and non-clinical university employees surveyed 4 – 5 weeks after work at home policies were implemented for those performing work deemed “non-essential” during the crisis phase of the pandemic. These findings uniquely highlight the associations of health and well-being with additional personal and work factors beyond those addressed in existing studies of HCW during the SARS-Cov-2 pandemic. Importantly, our study also reports on workers outside of clinical medicine, whose health and well-being has been minimally studied. A unique finding of this study is that the factors with the strongest consistent associations with all health and well-being outcomes in both clinical and non-clinical workers were items from the Family Supportive Supervisor Behavior Short-Form (FSSB-SF), a measure of general perception of family specific supervisory support,^8^ and a sum of eight stressors related to family/home life and financial security. Perceived supervisor support for family is a pathway through which employees develop perceptions of organizational support,^14^ plays a major role influencing the health and well-being of workers,^15^ and is associated with reduction in work-family conflict, improved well-being, and increased job satisfaction.^14,16^ Importantly, these factors are potentially modifiable by employer policies and practices.

Limitations of this study include its cross-sectional design, so associations between potential risk factors and health and well-being outcomes may not be causal. The overall response rate of 34.5% means that the respondents may not be fully representative of all university employees. Since the survey was anonymous, our study relies entirely on self-reported data. We studied employees of one university, who may not be representative of other workforces. The St. Louis region was an early adopter of physical distancing and has had a later peak of SARS-CoV-2 and a lower incidence of COVID-19 patients than some other areas of the US. Strengths of the study include its large size, examination of employees who are not in health care, and evaluation of both family/home stressors and workplace factors including supervisor support. To our knowledge this is the first large American study of mental health and well-being outcomes related to the pandemic outside of HCW. We will conduct repeated surveys over time to track changes in individual health and well-being over time, and to allow more robust causal inferences.

Our findings among clinical workers, both faculty (primarily physicians) and staff (primarily nurses) are broadly consistent with findings from other cross-sectional studies of HCW caring for COVID patients. A study of 1257 HCW in China^1^ used different instruments and found higher prevalence of depression and anxiety than seen in our study. Their study reported that HCW directly involved in the care of patients with COVID-19 were at a greater risk of anxiety and depression, similar to our findings of increased risks of stress, anxiety, burnout, and work exhaustion. A study of 906 HCW in Singapore and India,^17^ using the DASS-21, found moderate to severe stress in 3.8%, anxiety in 2.2%, and depression in 8.7%, much lower than the prevalence of 14.5%, 17.2%, and 16.4% seen in our study. Our finding that family/home stressors and supervisor support for family-work balance were strongly associated with mental health and well-being outcomes are consistent with the findings of a recent review^18^ of psychological reactions of HCW during past epidemics. Their analyses showed that responsibilities of caring for family members and lower household income were associated with poorer mental health outcomes among HCW. While HCW caring for COVID-19 patients had worse mental well-being than their fellow faculty and staff, those working from home or onsite in non-clinical roles also had appreciable rates of poor outcomes. While we do not have baseline measures for the well-being and mental health outcomes in our study, respondents described altered well-being related to COVID-19-related work/life changes, with 14.6% reporting “much worse” and 68% reported “much worse” or “somewhat worse” mental well-being. These findings are strikingly similar to those of an April 2020 poll by the Kaiser Family Foundation. Among those who had not experienced job or income loss, 15% reported major negative impacts on their mental health from worry or stress over coronavirus, and 54% reported some negative mental health impacts.^19^

University staff and to some extent faculty are representative of the larger non-clinical workforce that is undergoing uniquely stressful circumstances that blur the boundaries between work and family as people work from home, find it difficult to work because their children’s schools and daycares are closed, or worry about bringing an infection home to their families. While front-line HCW are at uniquely high risk due to their work, our study shows that effects of family and home stresses and of supervisor support play a large role in their health and well-being. Appreciation of these factors has been largely missing from studies of risk factors for mental health and well-being among HCW during this pandemic. These same family and home stresses and supervisor support also influence the health of the broader working population. As the pandemic continues in the months and perhaps years to come, our concern over the mental health and well-being of healthcare workers must broaden to include other worker groups as well.

There are many possible interventions to address the health and well-being of the clinical and non-clinical workforces. A systematic review found that organizational and social support, clear communication, and having a sense of control were protective factors for adverse mental health outcomes among healthcare workers during prior epidemics.^20^ Recent publications have stressed the importance of robust organizational responses to address the mental health and well-being of front-line HCW.^5,21^ Many of these interventions should be applicable outside of the healthcare setting. While interventions aimed at improving resilience among individual workers may lead to improvements in burnout and other well-being measures, organizational level interventions that reduce perceived work demands or increase resources are generally more effective.^22^ Our data would suggest that organizations should explicitly focus on improving supervisor support for work-family issues. Evaluation of interventions training supervisors in family supportive behaviors, including a study in healthcare workers, have suggested that such training is associated with improved reports of physical health, job satisfaction, job engagement, and decreased intent to leave the current job.^23,24^ Future research should include longitudinal studies to follow mental well-being over time, should include more workers outside of health care to better understand the effects on the broader population, and should test both individual level and institutional level interventions to mitigate the effects of the pandemic on mental health.

## Conclusions

Both health care and other workers have encountered worsened mental health and well-being as a result of the SARS-Cov-2 pandemic. Employers, health care systems, and public health agencies should begin interventions to improve mental health and overall well-being among HCW and the broader workforce. In addition to traditional wellness interventions addressing resilience and mental health issues among individual workers, responses should include support for work/family balance and other organizational changes to improve work conditions for health care and other workers.

## Data Availability

Anonymous survey data may be made available to interested researchers who provide written request to Dr. Evanoff at.

## Author Contributions

BAE was responsible for study conception and design, survey development, analytic oversight, and drafting of this article. JRS contributed to study conception and design, survey development, coordinating data collection, and provided input on analytic decisions, interpretation, and manuscript development. AMD provided input on study design, analytic decisions, interpretation of results, and manuscript development. LH contributed to survey development, managed online survey collection, conducted analyses, and assisted with manuscript development. EP assisted with study design, survey development, and data collection, contributed to development of the manuscript, and communicated results back to university employees. JGD, TK, and DLG provided input on study design, survey development, analytic decisions, interpretation of results, and manuscript development. All authors reviewed and approved the final manuscript draft.

